# CoVid-19: The Second Wave is not due to Cooling-down in Autumn

**DOI:** 10.1101/2020.11.27.20239772

**Authors:** Walter Langel

## Abstract

In analogy to influenza the second wave of the CoVid-19 disease is generally considered as being triggered by cooling-down in autumn and enhanced aerosol distribution. Here, the time histories of the total case numbers in three European states are quantitatively compared with those of Argentina by a generally applicable fit procedure. It turns out that Argentina on the southern hemisphere sees the second wave simultaneously with similar parameters as Europe. This discards the assumption of the influence of atmospheric cooling in winter and puts into question present models of SARS-CoV-2 spreading.

## 1 Introduction

The case numbers of the CoVid-19 disease are of worldwide interest and large effort is made to understand infection mechanisms and reduce infection rates. The person to person SARS-CoV-2 transfer by aerosols is considered to be dominant. According to general assumptions, this mechanism is enhanced when weather is cooling down and people spend more time inside closed buildings (1). The rapid increase of the infection rates in many countries since October 2020, addressed as “second wave”, is attributed to increasing aerosol transfer similarly to observations on influenza (2).

Data analysis proceeds often via compartment models which define different groups such as susceptible (S), infected (I) and recovered (R) individuals, and others. The flow between these groups is governed by equations analogous to chemical kinetics, mostly for first “reaction” order. Only the infection rate is proportional to the product of infected and susceptible, and the respective equation (eq (1) in (3)) is a second order process. This model describes outbreaks of a virus where the number I_0_ of infected persons is small at the beginning, and finally affects the full group of susceptible persons S.

The comparison of different countries and various outbreaks in these countries has to proceed via few significant free parameters, and here a modified SIR model is applied. The total case number in each country is decomposed into a sum of outbreaks N, and each outbreak is fitted by a single logistic function, which describes the infection of only a part S_N_ of the total population of this country by I_0,N_ of initially infected persons.

It was shown earlier that a similar approach can be applied to many states (4). Here, France, Germany, Spain and Argentina are considered: Germany and France have similar starting conditions, and Spain as a third European state has a significantly warmer climate, especially different in autumn from Germany, and thus should be less affected by the second wave. Argentina on the southern hemisphere goes in October and November from winter to summer while the three European countries see cooling from summer to winter. Here it is claimed that data from countries on the southern and northern hemisphere are very similar, and that this is not consistent with the omnipresent assumption of enhanced virus spreading in colder seasons.

## 2 Method

Each outbreak N is fitted by a single logistic function, as described earlier in (4):

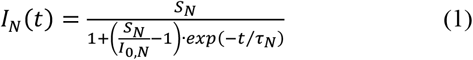

This well-known expression is the solution of the simple compartment model SIR (3). It is adapted to the observed case numbers and infection rates from very different countries by varying only a few parameters (Table 1):

**Table 1:**
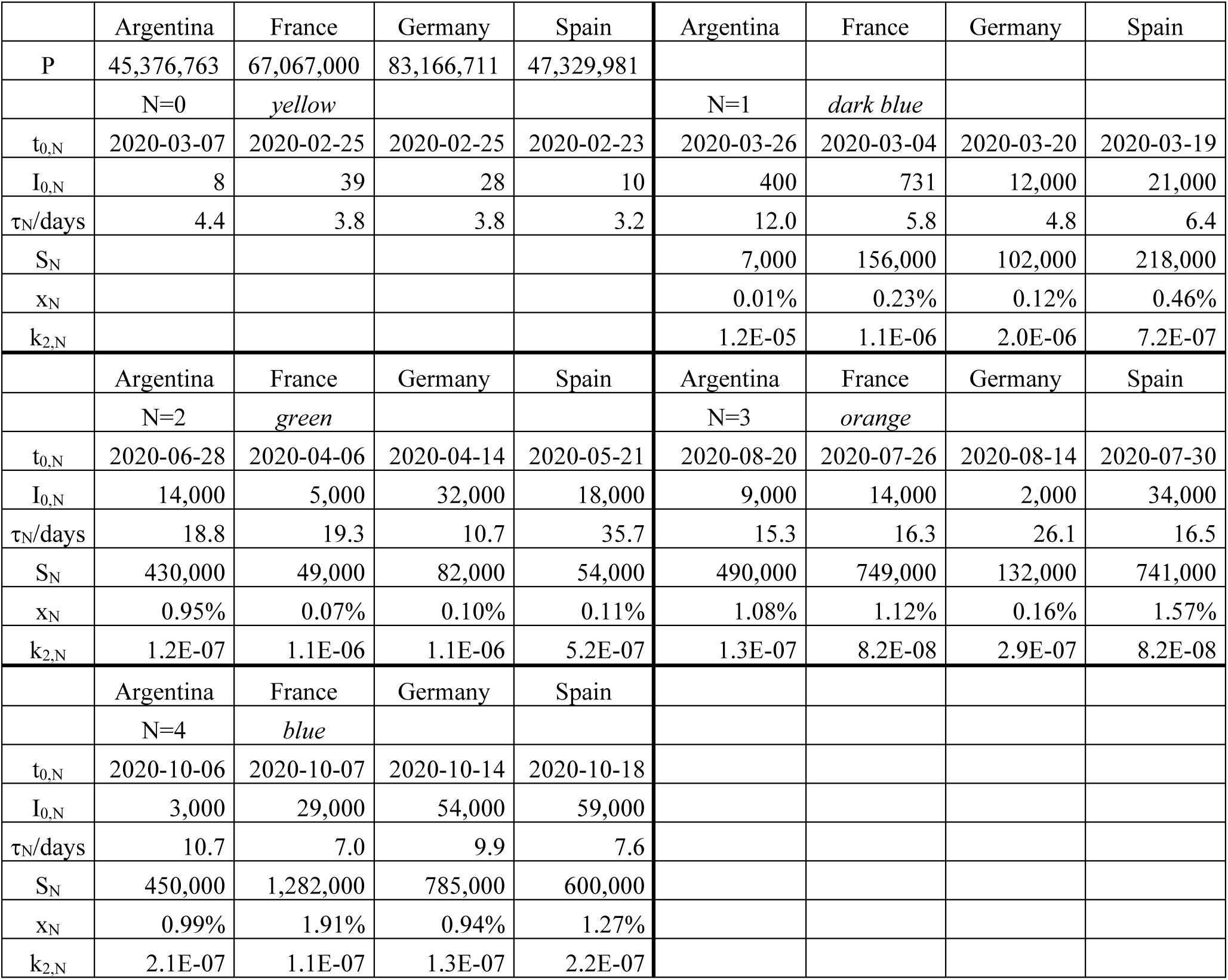
Fit parameters for CoVid-19 outbreaks according to eq. (2). The meaning of the parameters is explained in the text. For clarity, the numbers N of the outbreaks and the respective colors in Figures 1 and 2 are indicated. Outbreak N=0 was interrupted in the exponential phase, and thus S_0_ and the values of k_2,0_ and τ_0_ derived from it are not defined.

1) *I*_0,*N*_ ≪ *S*_*N*_ is the number of individuals triggering a new outbreak N.

2) The time constant *τ*_*N*_ of usually a few days has two implications:

- During the first days an exponential increase of both case number and infection rate is seen, and the often cited doubling time is related to *τ*_*N*_ by *τ*_2,*N*_ = *τ*_*N*_ *ln*(2).

- Later, increase is significantly slower than exponential and the case number and infection rate asymptotically attain a constant value *S*_*N*_ and zero, respectively. The time scale of the outbreak is also determined by *τ*_*N*_.

3) *S*_*N*_ is the maximum number of infected persons at infinite time in a given outbreak, i.e. the whole group, which is in reach of the initially infected *I*_0,*N*_. In the simplest case, *S*_*N*_ is as high as the total population P of the respective country (cf. (5)). Social distancing limits the total number of individuals, which may be attained by the initially infected group. This, in the modified SIR-model, *S*_*N*_ in each outbreak is much smaller than P, and *x*_*N*_ = *S*_*N*_ */P* ≪ 1. The second order rate constant *k*_2,*N*_ of the SIR model (cf. *λ* in (3)) is given as 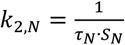 (in days^-1^susceptible persons^-1^).

Each outbreak N is described independently by a new logistic function *I*_*N*_(*t*), and distinct outbreaks of the virus are assigned to the observed data. Previously (4), outbreaks were considered separately in single time intervals. A first outbreak, N=0, showed purely exponential increase and was quickly interrupted after a few days. Sufficient data are now available for consistent fits over the whole time from end of January to mid-November. It thus turned out that the further outbreaks N=1-4 do not occur consecutively, but may also overlap on the time scale, and that the case numbers *I*_*N*_(*t*) add up to the total case number *I*(*t*) :

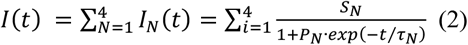

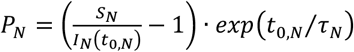 is used as fit parameter, and the number of infected persons triggering the start of the wave N is estimated as 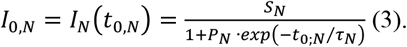

4) The additional parameter *t*_0,*N*_ is introduced since the outbreaks start at different times. It is determined as the date, when the observed total case number and infection rate significantly increase with respect to the respective calculated sums of the previous waves 1..N-1.

## 3 Results and Discussion

The observed total case numbers yield fairly smooth curves (Figure 1), and in many cases steps indicating a new outbreak are clearly visible. Even though a very simple model with few parameters (table 1) is underlying, the observed data are well fitted by equation (2). It is possible to extrapolate the curves over short times. By early identifying the deviation of observed data from the fit of the previous outbreaks (1..N-1), a spontaneous new outbreak N can be predicted.

**Figure 1:**
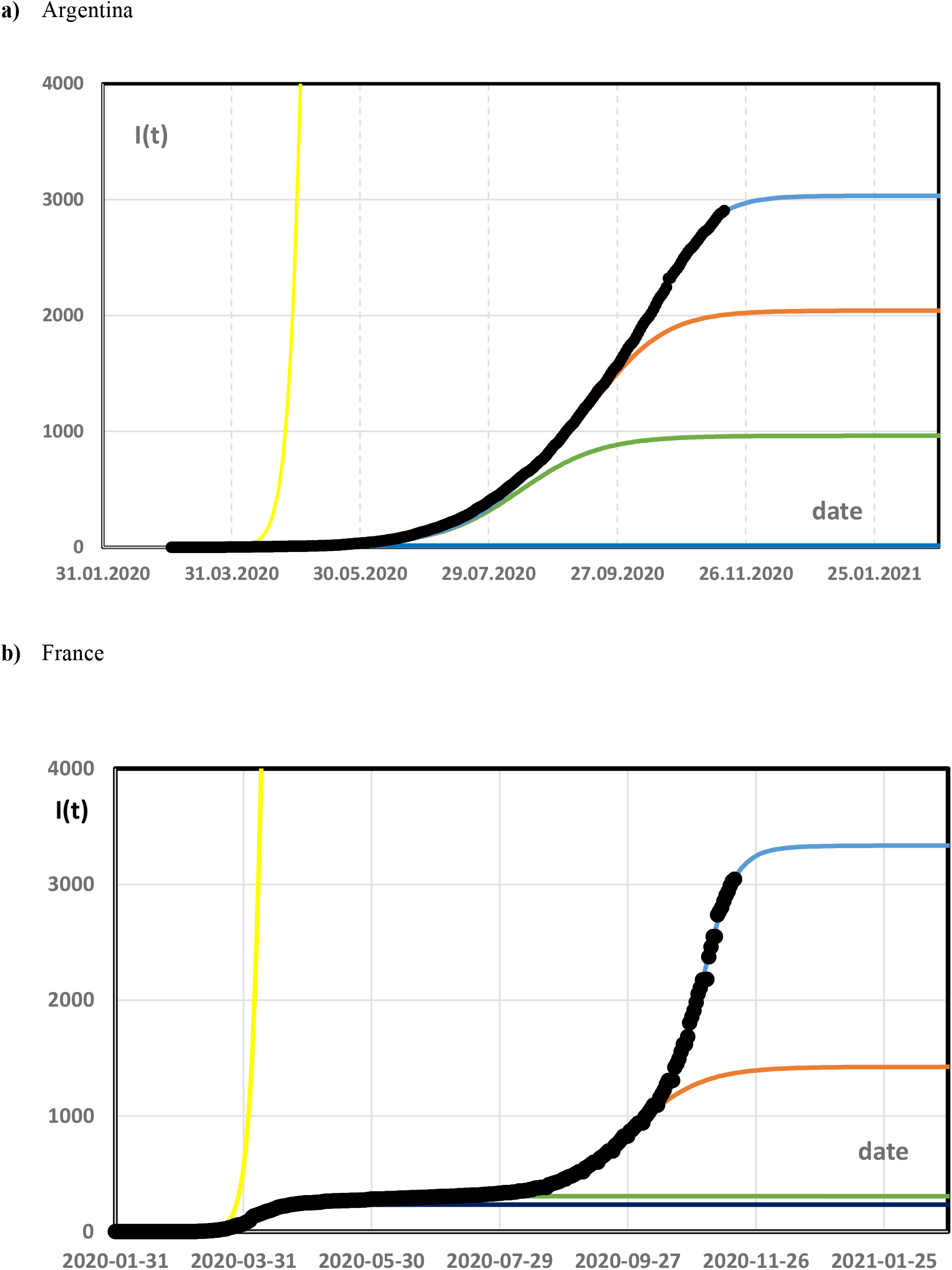
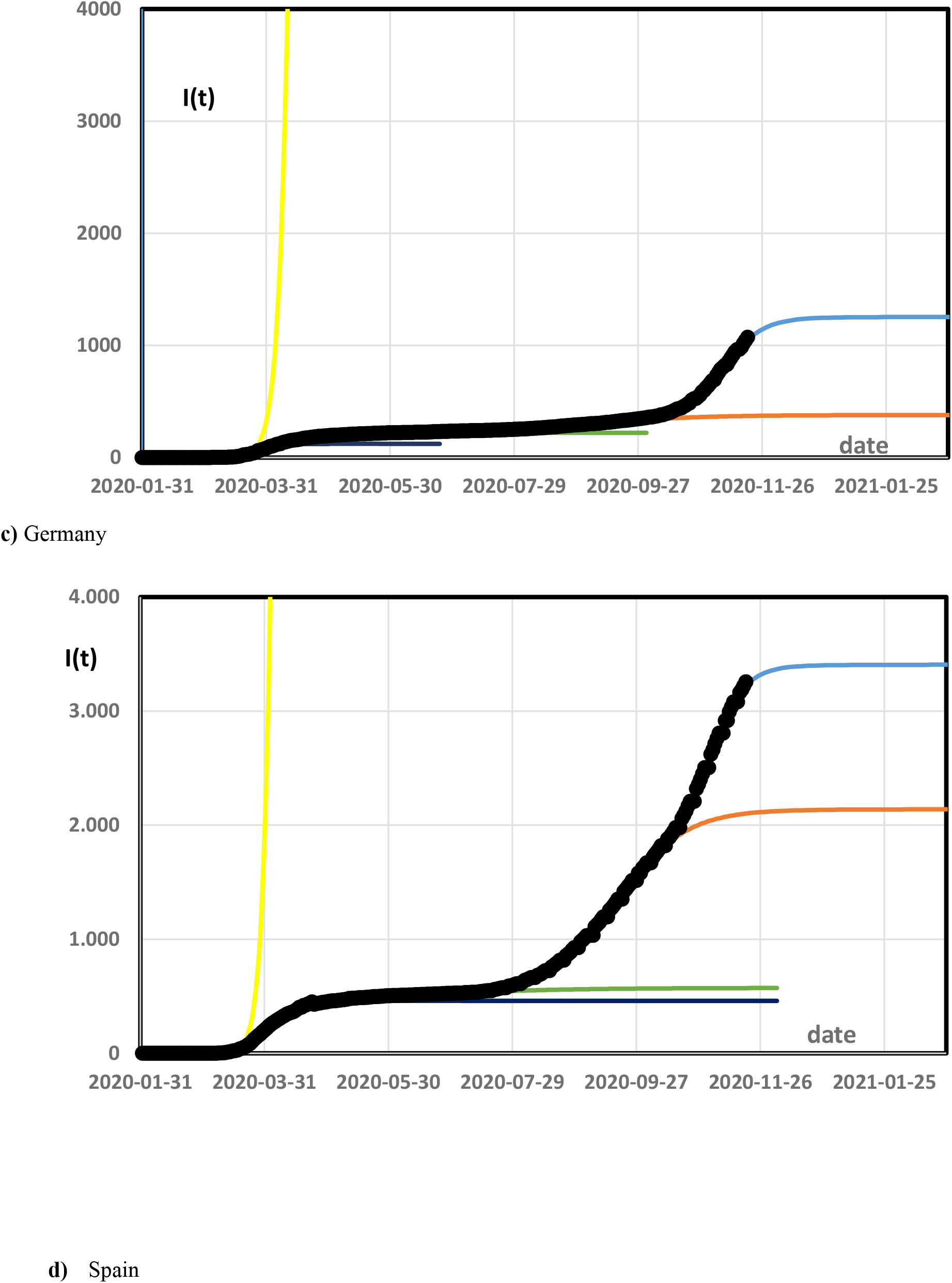
Total case numbers as a function of time (incidences per 100,000) are plotted. Dotted black: observed data ((8), (9)), yellow: extrapolated infection rate without social distancing (N=0), dark blue: small outbreak after the lock down (N=1), green: outbreak during release N=2), orange: enhanced outbreak in summer/winter (N=3), blue: enhanced second wave in autumn/spring since October (N=4).

Daily infection rates oscillate with a seven days period and are subjected to strong noise due to fluctuations in data collection (Figure 2). Calculating the day to day differences of this rate effectively yields the second derivative of the total case number, which is an extremely rough function. In media presentation the problem of weekly fluctuation is reduced either by showing a seven days running average, or by evaluating rate differences not between consecutive days, but between a day and the same weekday in the week before. In any case statistical noise is obscured by the weekly oscillations. The procedure adopted here circumvents this problem by fitting the rather smooth total case number by an analytic function and then evaluating the infection rate as a smooth curve from the fit. The infection rate is essentially the first derivative of the total case number as a function of time, and shows a bell shaped structure for each outbreak.

**Figure 2:**
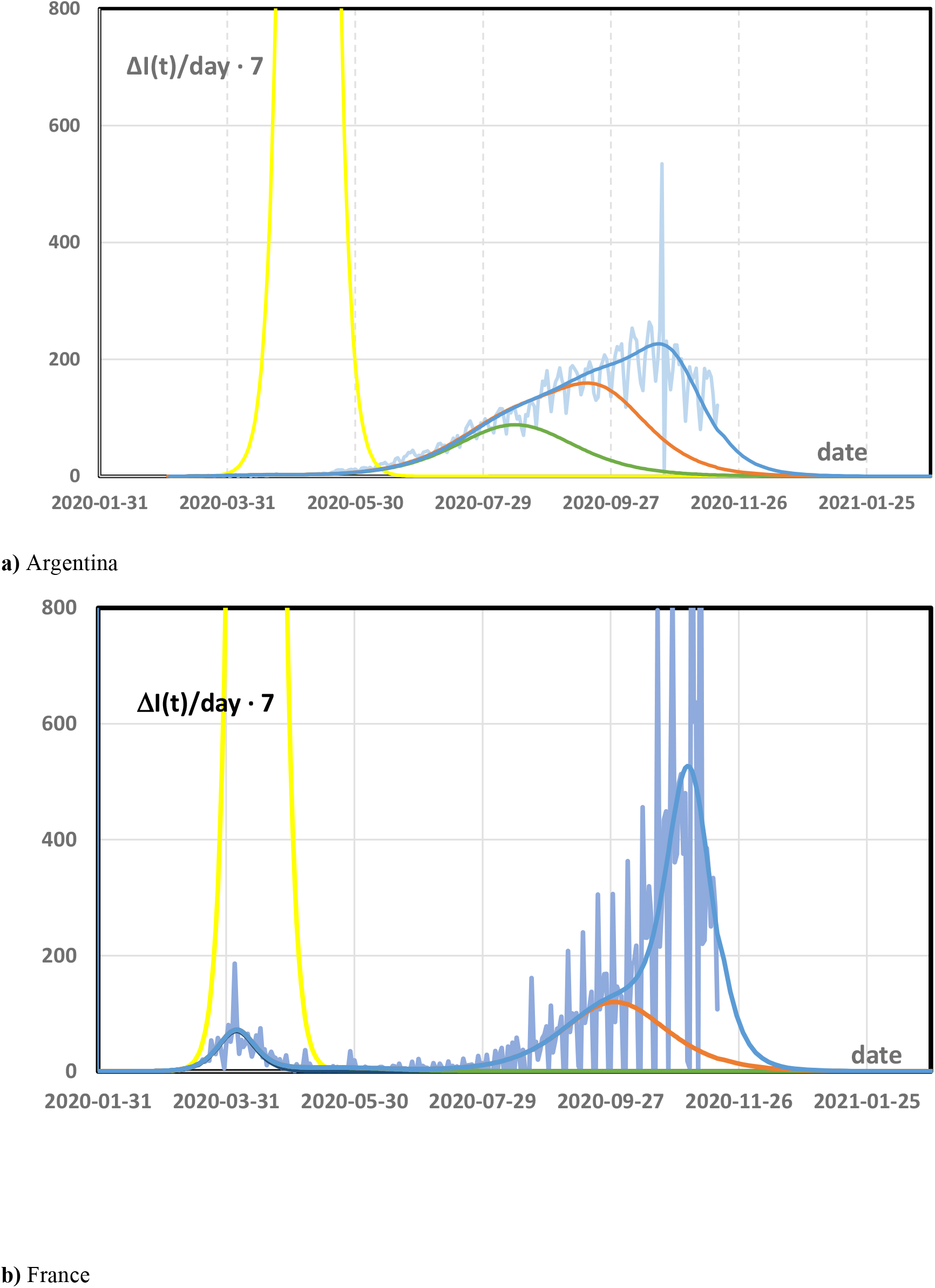
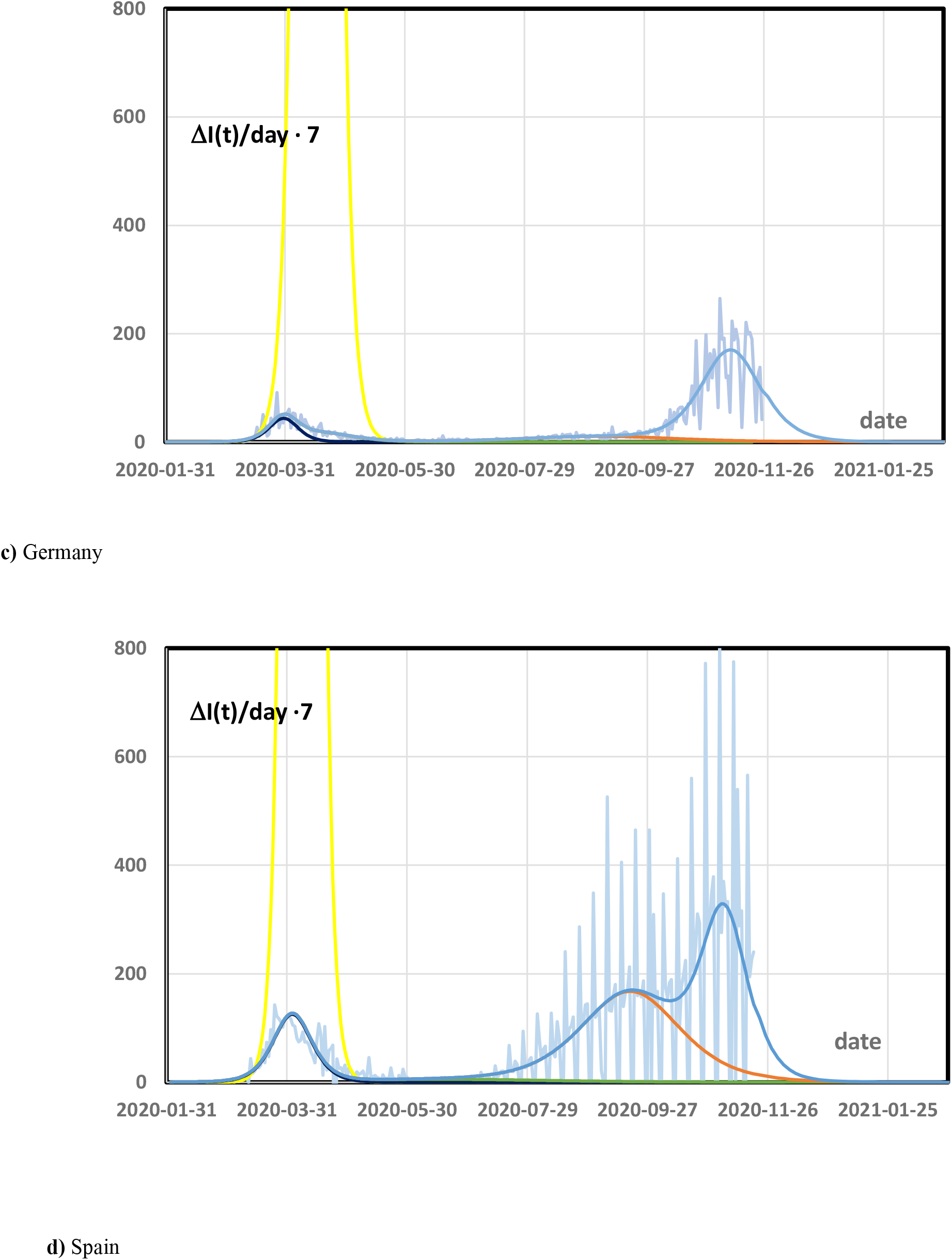
Infection rate per 100,000 persons per week. The vertical axis directly compares with the numbers for incidences per 100,000 persons per week as used e.g. for identifying high risk regions. Light blue: observed infection rates with strong weekly oscillations, colors for fitted outbreaks as in Figure 1. All infection rates were calculated from the respective data for total case numbers by taking the day-by-day differences and multiplying by seven.

Figures 1 and 2 and table 1 show plots and parameters for fitting the observed data by so far five outbreaks:

(1) N=0: The strictly exponential increase was interrupted after a few weeks in March by any form of social distancing in all countries before a significant deviation from the exponential increase or a saturation were recognizable. I_0,0_ was very low, but infection could have gone to very high numbers of *S*_0_ *≈ P* as provided in the original SIR-approach. The time constants were 3-4 days, the doubling time then only 2.3-3 days.

The following four outbreaks all took place under conditions of social distancing. A few weeks after the beginning t_0,N_, the additional case number I_N_(t) clearly drops below the initially exponential increase (4). The time τ_N_ is significantly higher for N=1-4 than for N=0, and the outbreaks continue independently:

(2) N=1: The first short outbreak after lock down or other measures started end of February or beginning of March 2020, and affected only a few tenth of percent of the population in Europe, whereas S_1_ was even nearly negligible in Argentina (cf. Fig 1a). Without the sequence of continuing spontaneous outbreaks, the pandemic should have ended in June 2020 (6). From the point of kinetics, this outbreak still had rather short time constants and high rate constants of around six days and 10^−6^, respectively.

(3) N=2: The N=1 outbreak was immediately followed by a next one starting April to June. The case numbers in this time were often ascribed to a release of the Anti-Corona measures. The initial number of infected people was significant (I_0,2_ of a 5-30 thousand), but the total number of infections only moderately increased by about 0.1% of the population in Europe. Only in Argentina, which had seen small impact of the N=1 outbreak, the case number was higher, around 1%. The time scale of this outbreak is considerably longer than of the previous one (τ_2,N_ around 20 days), but the rate constant is still of the order of 10^−6^ in Europe.

(4) N=3, 4: After a calm period in May and June, two severe outbreaks were observed in late summer/winter and early autumn/spring without having an obvious origin. Both overlap each other in a similar way in Argentina, France and Spain (Figures 2a, c, d). In Germany the outbreak N=3 had small impact, (x_3_=0.2%), and therefore the rapid increase of infection rate in October became very obvious. These two outbreaks together are considered here as a quantitative description of the “second wave”.

The numbers of initially infected persons of I_0,3_ and I_0,4_ are in a similar range of 2-50.000. The final values S_3_ and S_4_ during the second wave are significantly higher than S_1_, S_2_ immediately after the first measures in March. Thus, x_3_ and x_4_ are around 1-2% of the total population, and add up to an infection of 1.1-3% of the respective total population.

All rate constants k_2,3_ and k_2,4_ were close to 10^−7^ and thus significantly lower than for N=1 or 2. It may be speculated that this indicates people being more cautious than in the beginning of the pandemic. The outbreak N=3 started end of July till mid-August in the four considered countries with a similar time scale of τ_3_ around 15-25 days as N=2. N=4 started in the first half of October even more synchronously in the four countries than the previous outbreaks. It has a smaller time scale with τ_4_ around 10 days and contributed mainly to the rapidly increasing high infection rates and incidences in October and November 2020.

## 4 Conclusion

A thorough quantitative analysis shows that the total case numbers and infection rates are not only described as first and second wave, but that a total of five outbreaks has to be distinguished so far. They typically start with a small number of infected persons, in the beginning few 100, later with in increasing number I_0,N_ up to 50.000. During the infection process, the total case numbers typically increase by a factor of ten; but the ratio S_N_/I_0,N_ varies in a large range, as I_0,N_ is not very strictly defined by the fit procedure.

In many cases, new outbreaks start parallel to previous ones, while these still generate significant infection rates. The spontaneous appearance of new infection outbreaks with high initial case numbers shows that measures such as the lock down earlier in 2020 had had no definite success.

In large countries outbreaks from different regions could be shifted with respect to each other, and their superposition could scramble the data to be fitted. It has been shown in (4) that this is not the case. Obviously the outbreaks are correlated even under conditions of reduced personal exchange. Thus, new outbreaks start spontaneously, but simultaneously in different countries.

The fits (Figure 1) yield a fairly stable prognosis as long as only existing breakouts are continued. From the noisy infection rate data, this is possible alone only with large computational effort, if at all (7). Prognosis on a longer timescale and for the end of the infection spreading is not possible due to spontaneous outbreaks.

Here, case numbers and infection rates of Argentina were compared with respective data from Europe. The main point to make is that the second wave occurs in a country on the southern hemisphere on a quite similar time scale and with similar incidences as in European countries of comparable sizes. As Argentina now sees the transition from winter to summer rather than from summer to winter as in Europe, this similarity discards the common theory that the second wave is due to autumn cooling. Moreover, the second wave should have larger impact in Germany than in Spain, where the climate in autumn still is rather mild. The opposite is observed, the country is far more severely hit than Germany. These observations leads to the speculative question, if the person to person transfer of the SARS-CoV-2 virus in aerosols is indeed the only efficient infection mechanism. As nearly all measures in most countries aim at reducing the aerosol mechanism, bypassing it by another infection channel would explain, why the success of many anti-Corona measures is limited.

## Data Availability

Data input comes from publically available Corona case numbers.

## References

1. Werner C, Friebe R, Eickmeier P. So ließe sich eine Massenansteckung in der kalten Jahreszeit verhindern (German). [Online].; 2020 [cited 2020 November 27]. Available from: https://www.tagesspiegel.de/wissen/corona-herbst-mit-zweiter-welle-so-liesse-sich-eine-massenansteckung-in-der-kalten-jahreszeit-verhindern/26099728.html.

2. Foster H. The Reason for the Season: why flu strikes in winter. [Online].; 2014 [cited 2020 November 27]. Available from:]http://sitn.hms.harvard.edu/flash/2014/the-reason-for-the-season-why-flu-strikes-in-winter/.

3. Amaro JE, Dudouet J, Orce JN. Global analysis of the COVID-19 pandemic using simple epidemiological models. Appl. Math, Mod. 2021: p. 995–1008.

4. Langel W. medrXiv. [Online].; 2020 [cited 2020 November 26]. Available from: https://doi.org/10.1101/2020.06.17.20134254.

5. wikipedia: List of countries and dependencies by population. [Online]. [cited 2020 June 18]. Available from: https://en.wikipedia.org/wiki/List_of_countries_and_dependencies_by_population.

6. Langel W. Extrapolation of infection data for the CoVid-19 virus and estimate of the pandemic time scale. medRxiv [Internet].; 2020 [cited 2020 June 19]. Available from: https://doi.org/10.1101/2020.06.17.20134254.

7. Pipa G. Bayessches räumlich-zeitliches Interaktionsmodell für Covid-19 (German). [Online].; 2020 [cited 2020 November 27]. Available from: https://covid19-bayesian.fz-juelich.de/.

8. Radtke R. Fallzahl des Coronavirus (COVID-19) in Deutschland, Frankreich und Spanien 2020 (German). [Online].; 2020 [cited 2020 November 27] [data from the John Hopkins institute, USA]. Available from: https://de.statista.com/statistik/daten/studie/1101414/umfrage/fallzahl-des-coronavirus-in-deutschland-frankreich-spanien/#professional.

9. worldometer. [Online].; 2020 [cited 2020 November 17]. Available from: https://www.worldometers.info/coronavirus/country/argentina/.

